# Applying causal inference and Bayesian statistics to understanding vaccine safety signals — a simulation study

**DOI:** 10.1101/2024.03.03.24303687

**Authors:** Evelyn Tay, Michael Dymock, Laura Lopez, Catherine Glover, Yuanfei Anny Huang, K. Shuvo Bakar, Thomas Snelling, Julie A. Marsh, Yue Wu

**Affiliations:** Wesfarmers Centre of Vaccines and Infectious Diseases, Telethon Kids Institute, Nedlands, Western Australia, Australia; National Centre for Immunisation Research and Surveillance, Westmead, New South Wales, Australia; School of Public Health, University of Sydney, Sydney, New South Wales, Australia; Sydney Children’s Hospital Network, Westmead, New South Wales, Australia; Centre for Child Health Research, University of Western Australia, Nedlands, Western Australia, Australia

## Abstract

Community perception of the safety of vaccines in the interest of public health influences vaccine uptake. Challenges for active vaccine safety monitoring include survey response rates, unbiased reporting and the balance between specificity and sensitivity of signal detection methods. To address these problems, we used causal DAGs and statistical methods to guide understanding of biological and behavioural factors which may influence vaccine safety signal detection. The DAGs informed the generation of scenarios in which these factors were varied. A posterior predictive analysis (PPA) signal detection method, based upon a Bayesian logistic model, was used to detect signals across the scenarios. In the high probability of severe reaction scenarios, true signals were generated where there was higher survey participation with more survey responder AEFI data available for analysis. In the low probability of severe reaction scenarios, false signals were generated when there was a strong influence of reaction severity on survey participation and reports of severe reactions. Low rates of survey participation reduce the amount of data available to inform the parameters of the statistical model, and therefore reduce the certainty regarding the value of these parameters. We obtained insights into the value of the causal DAG to account for survey non-response, to guide understanding of short-term vaccine safety, interpret the results of the PPA analysis under plausible scenarios, and review implications for future vaccine safety monitoring

## Introduction

Routinely administered vaccines in the interest of public health are safe, but mild reactions are common and serious reactions occasionally occur, even for vaccines with otherwise excellent safety profiles. In 2010, an influenza vaccine formulation was associated with an increased risk of febrile convulsions in Australian children under five years old [1]; this led to the discontinued use of the particular formulation in children, but not before a temporary suspension of all influenza vaccination in young children, affecting public confidence in childhood immunisation, which reduced vaccine coverage [2]. Low vaccine coverage increases the risk of infections caused by vaccine preventable diseases [3]. There is an ongoing need to monitor, detect and address potential vaccine safety issues as soon as possible after they emerge, which is essential to public confidence in vaccination.

Monitoring can be performed for either solicited (active) or unsolicited (passive) reports of adverse events following immunisation (AEFI), which are undesirable clinical events occurring after the administration of a vaccine, irrespective of whether any causal relationship with the vaccine exists [4]. AEFI are graded according to their severity [5] ranging from mild AEFI that do not interfere with a person’s activity to severe AEFI preventing normal activity and/or requiring medical attention. Australia’s active vaccine safety surveillance system AusVaxSafety monitors the frequency of solicited acute AEFI for vaccines delivered through the national immunisation program [6]. Since 2014, data has been collected through AEFI surveys sent to vaccine recipients via SMS or email three days after vaccination. Reports of seeking medical advice or attention are taken to be an indicator of severity, although health care seeking may also be influenced by vaccine concerns raised in the media and other procedural factors. The rate of survey-reported medically attended AEFI (MA) is monitored to identify potentially important vaccine safety issues that might require the suspension of a vaccine program pending detailed investigation. In 2021, AEFI surveillance for the newly developed and rapidly deployed Coronavirus 2019 (COVID-19) vaccines required extension of the AusVaxSafety post-vaccination surveys, including reports of any medical care or medical advice sought (attendance at a primary care clinic or emergency department or telehealth advice), the impact of reported AEFI on daily activities, and the presence of underlying health conditions [7].

A vaccine ‘safety signal’ is an unexpected (or unexpectedly frequent) association between a specific AEFI and a specific vaccine which requires investigation into whether a clinically important causal relationship exists [8]. To detect safety signals, AusVaxSafety employs a posterior predictive analysis (PPA) method; the PPA method is based on the posterior predicted distribution from a Bayesian logistic model adjusted for age, sex and co-morbidities [9]. The PPA method depends upon solicited reports of MAs; this, in turn, depends on survey participation which can vary across demographic groups and over time, leading to variable response rates. For example, over the first year of the Australian COVID-19 vaccine programme, the survey response rate declined from approximately 70% to 30% (unpublished data).

The challenge of vaccine safety monitoring is to identify sensitive and timely safety signals that unveil true vaccine safety issues, while minimising the frequency of false detections. To do this, survey-based vaccine safety monitoring systems must correctly interpret AEFI data from the subset of responders that exhibits non-random missingness. For example, responder rates differ between age and sex, and potentially between those who experience an AEFI and those who do not. Structured patterns of missing data may occur when vaccines are rolled out at different times to selected higher risk or specific age groups. A limitation of routine analytic approaches is the failure to properly account for these patterns of missingness in the data. The causal directed acyclic graphs (DAGs) are increasingly used to model data-generating processes in applied health research [10]. Causal DAGs depict and communicate one’s understanding of complex problem domains or hypotheses, which can subsequently guide the analysis and modelling assumptions [11]. For example, behavioural researchers have applied causal and statistical modelling techniques to understand the generalisability of samples for cross-cultural comparison [12]. In order to correctly infer the true AEFI rates among the vaccinated population from the survey responder data, some insight is required into the frequency of AEFI among survey non-responders.

In this paper, we approached this problem using causal DAGs created through expert elicitation to derive an assumed data generating process; from this, we simulated AusVaxSafety survey data collected under a range of scenarios. The synthetic scenarios modelled a range of biological and behavioural factors that plausibly influence the frequency of actual and reported medical attendance following immunisation (MA). We applied the PPA method to the simulated data under these scenarios and assessed its performance in flagging a safety signal for MA in survey responders in relation to the true rate of MA in both survey responders and non-responders in the simulated data. Using these methods we quantified how changes in important biological and behavioural factors could influence the reporting of MAs and affect the performance of signal detection using the PPA method. We obtained insights into the value of the causal DAG to account for survey non-response, to guide understanding of short-term vaccine safety, interpret the results of the PPA analysis under plausible scenarios, and review implications for future vaccine safety monitoring.

## Methods

### The scientific causal models

A directed acyclic graph (DAG) consists of nodes that represent random variables that may or may not be observed (in the form of data), and arrows (arcs) that indicate a possible direct influence of predecessor (or parent) variables on their child nodes (nodes extending from other nodes). A causal DAG is one in which the arcs are intended to represent influences that are causal and not just associative. In this study, the purpose of creating the causal DAGs was to represent how the AusVaxSafety active surveillance system for short-term AEFI monitoring operates as a complex system (the *full DAG*). It addresses how this complexity may affect the reporting of AEFI, especially MA, and how this may, in turn, influence the detection of safety signals. Using a previously published causal knowledge engineering process [13], the DAGs were drafted, refined, and applied for the stated purpose. Domain experts were consulted to advise on the selection of relevant variables, the causal structure, and the face validity of the final DAGs. The domain experts were public health practitioners, clinical vaccinologists, program managers familiar with AusVaxSafety’s data capture processes, and statisticians responsible for the analysis of the survey data and reporting of vaccine safety signals. From the full DAG, we extracted a *simplified DAG*, which preserved all causal assumptions from the full DAG.

### Data simulation and scenarios

To explore factors that influence signal detection via statistical analysis, we used all variables in the simplified DAG to generate complete data (i.e., without missingness) relevant to the problem domain, including for vaccine recipients who respond to the survey (observed) and those who do not. Binary discrete variables were sampled from Bernoulli distributions. Age and other continuous variables were sampled from Gaussian distributions and then categorised, e.g., ‘below 50 years old’ and ‘50 years old and above’. The choice of parameters for the statistical distributions used in simulation was informed by reasonable and plausible ranges. Guided by the causal DAGs, we designed hypothetical investigation scenarios of scientific interest that reflect potential variations in biological, behavioural and procedural assumptions, which may lead to distinct patterns of the reporting of MA. For each scenario, we generated 5,000 simulations to investigate the operating characteristics of the statistical signal detection method.

### The statistical signal detection method

We were interested in how a safety signal may be generated under each hypothetical scenario of interest using the PPA method. The PPA method identifies signals when the number of reported MAs in an investigation period exceeds a pre-specified threshold, defined as the 99^th^ percentile of the posterior predicted distribution for the number of reported MAs under a Bayesian statistical model, informed by (historic) reference data. The parameters of the Bayesian statistical model used to derive the threshold are regularly updated using survey data and account for age as one of the explanatory variables (as illustrated in Figure 1). We summarise the simulations for each investigation scenario in four ways: 1) the mean number of reported MAs, 2) the mean threshold value (from the posterior predictive distribution for the number of MAs), 3) the mean percentile of the number of reported MAs of the posterior predicted distribution (as an indication of how closely the mean number of reported MAs approaches the mean signal threshold), and 4) the proportion of simulations where the number of reported MAs exceeds the signal threshold, *i.e.*, the proportion of simulations in which a signal has been generated. This is illustrated in Figure 1.

**Figure 1:**
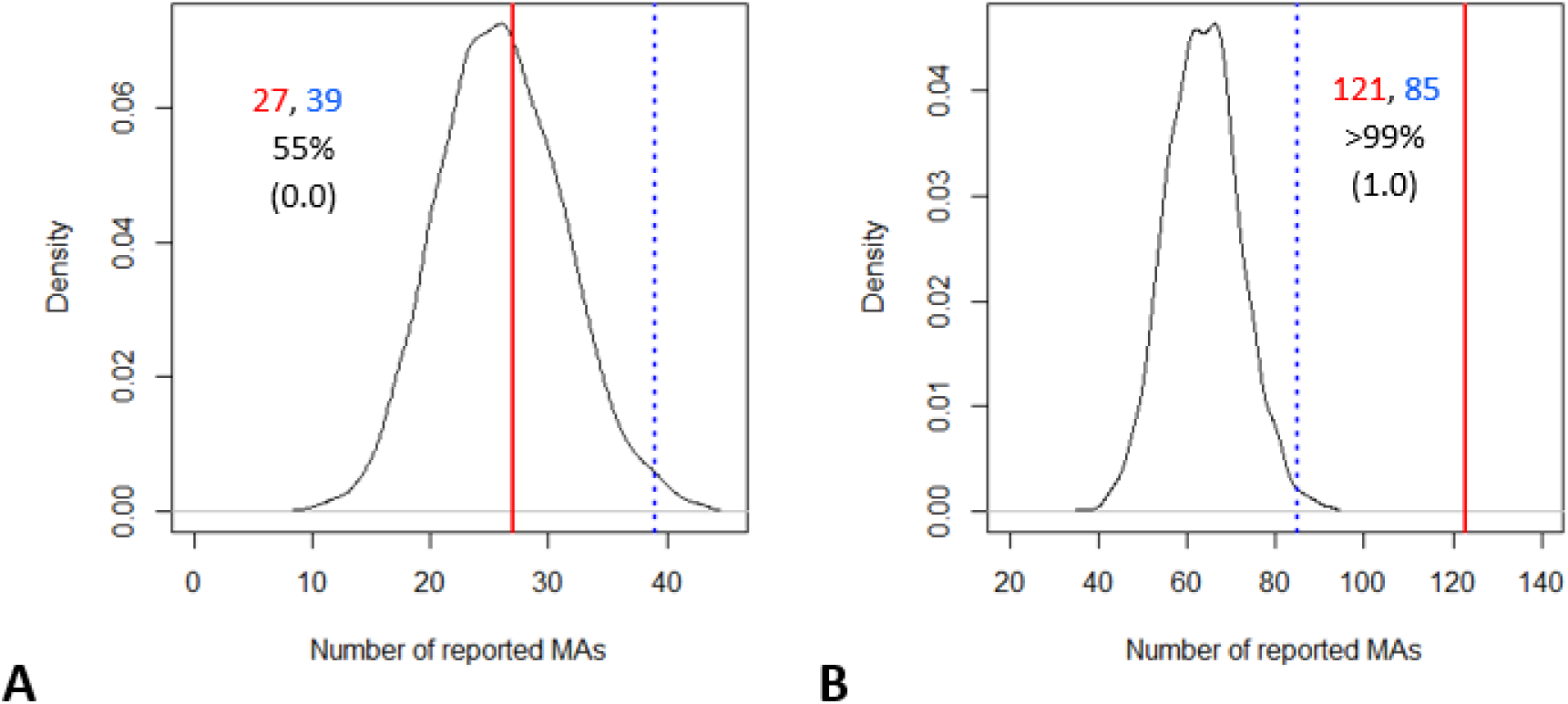
Illustrative outputs of the PPA method from a single simulation. The number of observed MAs, reported in red and the signal threshold reported in blue, are also indicated by the solid red lines and dotted blue lines, respectively. The percentile of the posterior predicted distribution is used here as an indication of distance of the number of reported MAs from the signal threshold. As the figure illustrates the outputs of just one simulation, the proportion of simulations with an alerted signal is 0.0 in Fig. 1A and 1.0 in Fig. 1B.

We considered a simplified version of the PPA consistent with our simplified DAG with only one explanatory variable, age, categorised into *<* 50 years old (denoted as *g* = 0) and *≥* 50 years old (i.e., *g* = 1). The number of MAs was modelled as arising from a Binomial distribution:

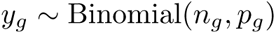

where:

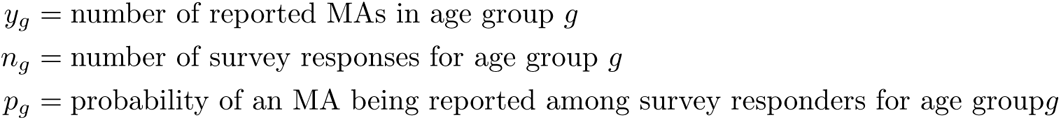

The linear predictor is:

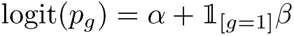

where:

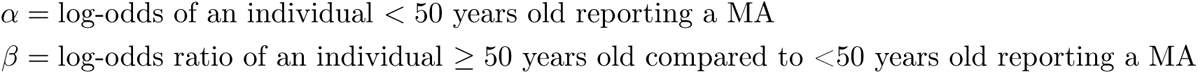

The model parameters were given the following weakly informative priors [9]:

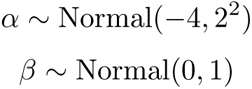

The prior distribution for *α* induces a 95% credible interval between approximately 0% and 48% for the probability of an individual *<* 50 years old reporting a MA. We used GeNIe software to build and depict the DAGs presented here [14]. All simulations and analytical programming were conducted in Stan [15] via the *cmdstanR* package [16] in the R statistical programming language v4.2.2. [17].Posterior distributions were estimated via Markov-chain Monte Carlo (MCMC) using stan’s Hamiltonian Monte Carlo algorithm with four MCMC chains, run in parallel, with warm-up and sampling phases each running for 1,000 iterations. The open source codes are available: www.github.com/ECSTay/AVSCausalModel

## Results

### Causal models

Figure 2 presents the full DAG which consists of 37 variables. Nine variables (blue nodes) depict the background risk factors for vaccine recipients. Six variables (green nodes) depict key events initiated in the health system (*e.g.*, the distribution of vaccines). The spectrum of expected AEFI is depicted by 7 variables (pink nodes), varying in expected frequency from common to rare (*e.g.*, 16% for fever [18] and 2.7 events out of 100,000 persons for chest pain [19], following vaccination with BNT162b2). Background variables and AEFI together drive the vaccine recipient’s perceived seriousness of an AEFI, which subsequently drives one’s overall level of concern regarding an AEFI. The level of concern can be further influenced by a recipient’s demographic background, which vaccine they received, and any contemporary factors that increase community concerns about vaccination in general, or the particular vaccine (*e.g.*, news about vaccine safety issues). The AEFI, its perceived seriousness, and the recipient’s level of concern, together drive the recipient’s actions including whether they seek medical attention and/or report the AEFI if surveyed. There are 11 actions (purple nodes) and 2 temporary factors (yellow nodes) modelled in the full DAG to illustrate the problem domain. See the Supplementary Dictionary for the definition of each variable and a detailed description of the DAG structure. At a high-level, there are three main processes that collectively culminate in the ascertainment of a MA report: 1) the vaccine reaction (biological process), 2) the seeking of medical attention (behavioural process), and 3) the data capture process (procedural process). There are many potential interactions among these processes. The key variables extracted to form the simplified DAG are highlighted in blue text.

**Figure 2:**
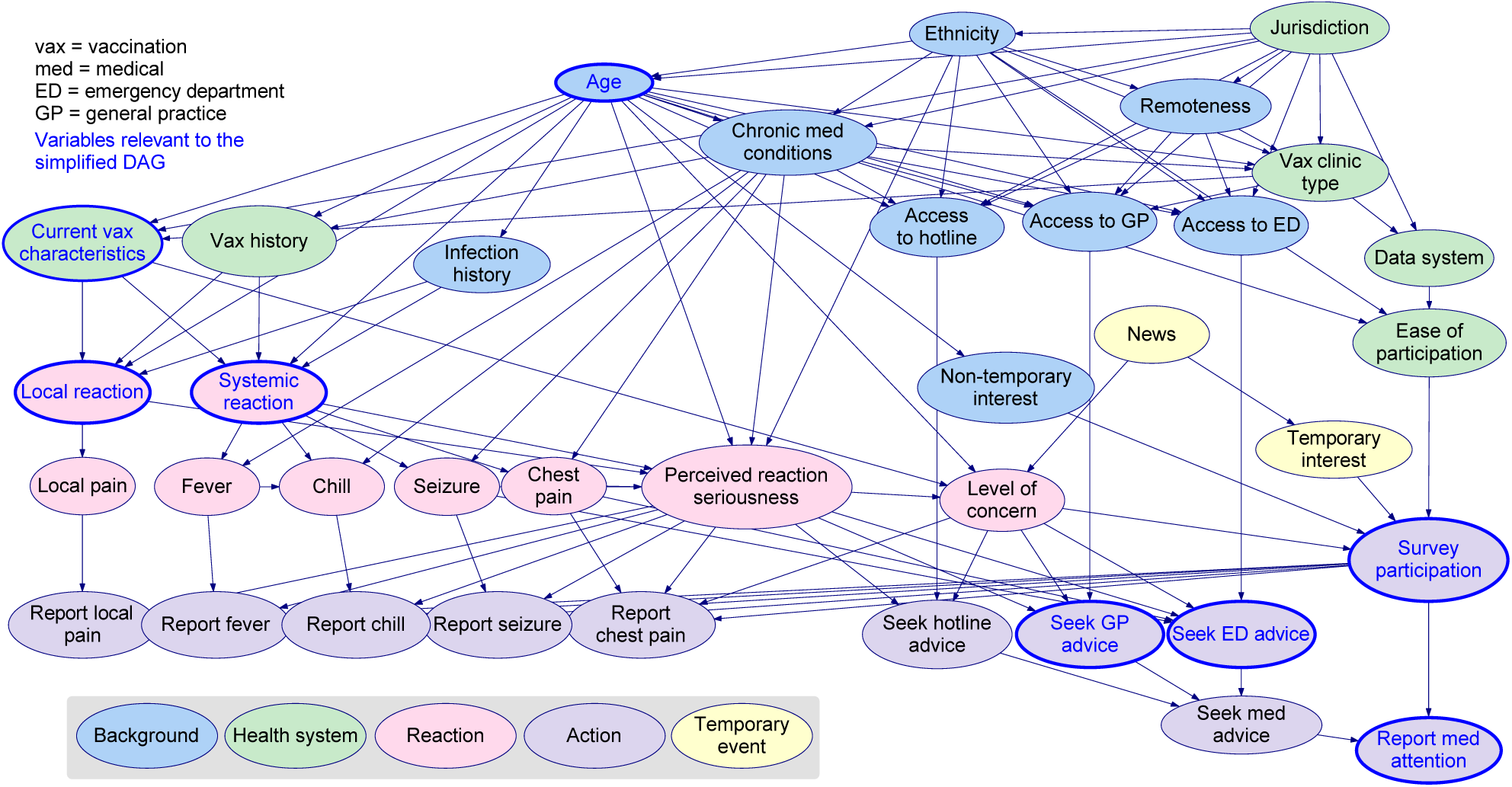
The full DAG which depicts how the AusVaxSafety active surveillance system for short-term AEFI monitoring operates as a complex system. See the Supplementary Dictionary for the definition of each variable and a detailed description of the DAG structure.

The simplified DAG (Figure 3) contains the key variables that we considered to adequately capture the full causal model for the purpose of subsequent investigations, namely the person’s age at vaccination (age), vaccine received (vaccine), whether they experience reaction within 3 days following vaccination (reaction), whether they seek medical attention (MA), whether they respond to, or participate in the survey (survey participation), and whether they report seeking medical attention (report MA). These represent a minimum set of variables required to address how biological, behavioural and procedural processes interact to influence the probability of a reported MA. In Section 3.2, we describe the design and underlying rationale for each scenario and the assumed data generation process. We assumed that the method of survey collection is constant and external influences are time invariant.

**Figure 3:**
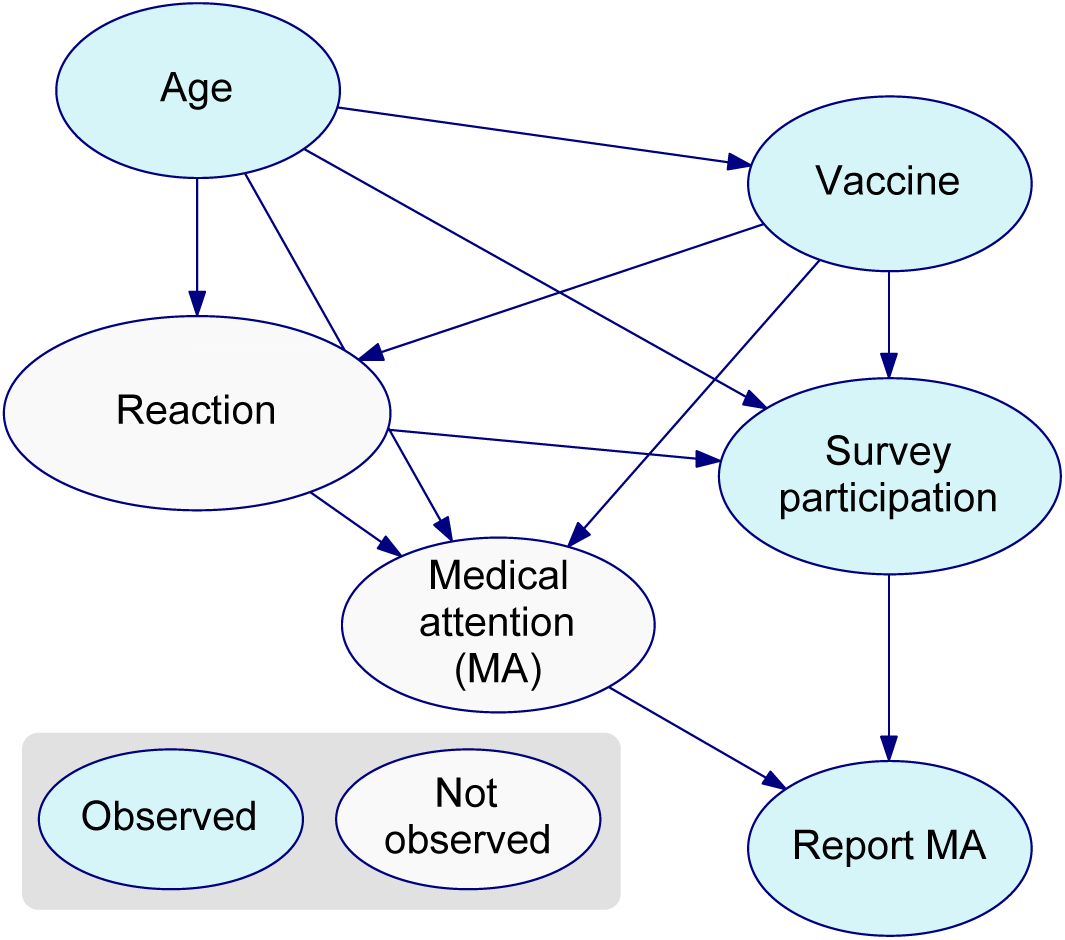
The simplified DAG. which depicts the relationship of a minimum set of variables required to investigate how biological, behavioural and procedural processes interact to influence the probability of a reported MA using PPA method. See the Table 1 for the definition of each variable, and see Supplementary Dictionary for how each variable is aligned with the full DAG.

**Table 1:**
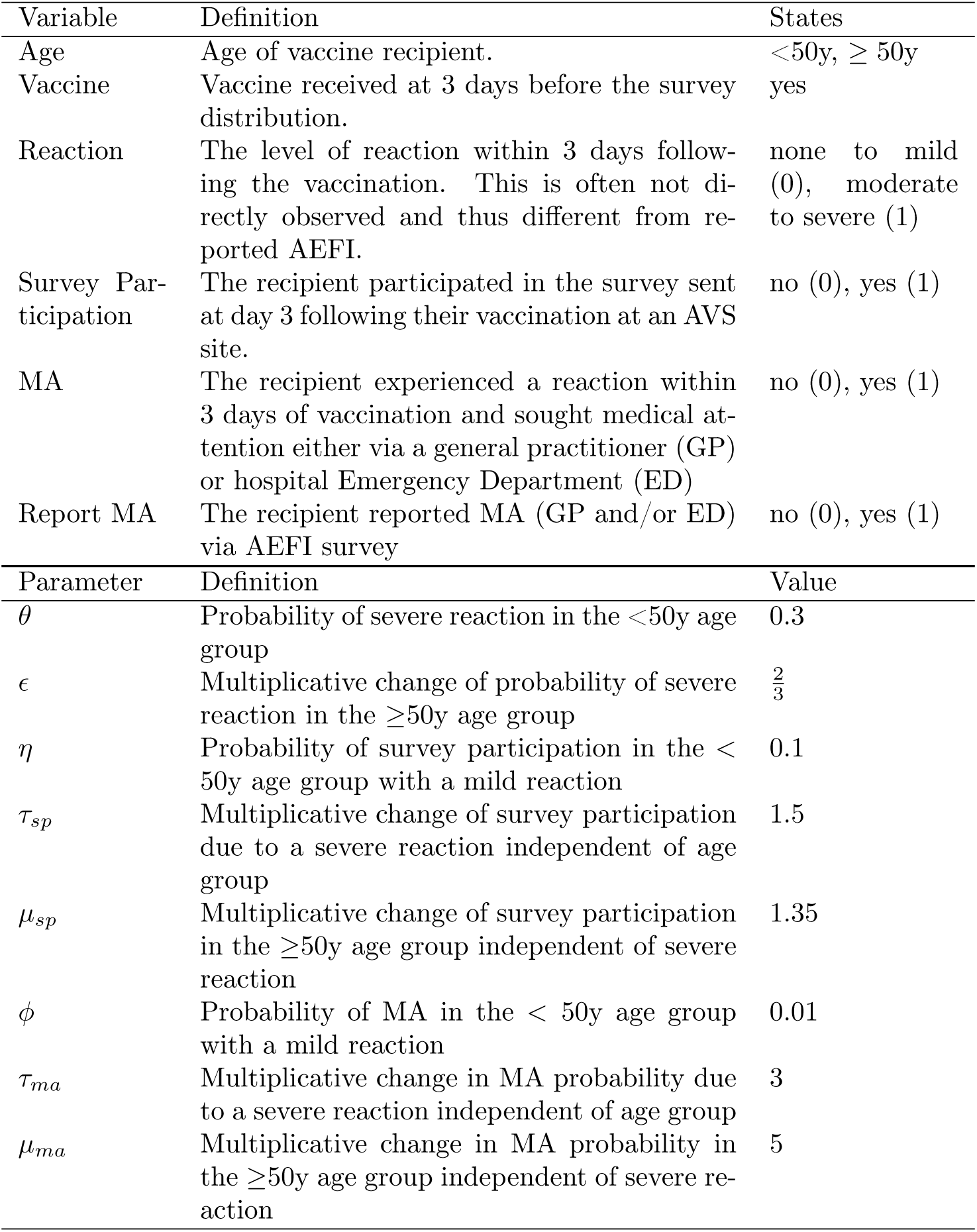
Simplified DAG variables and data simulation parameters.

### Data simulation and scenario design

We simulated the five variables represented in the simplified DAG using eight parameters as defined in Table 1. The vaccine variable was not explicitly included in the statistical model as a single type of hypothetical vaccine was considered. For survey responders, only age and reported MA can be observed and the other factors are not observed. In the simulations these parameters and their values were chosen based on several key assumptions. In brief, we assumed that the more severe an AEFI, the more likely a person is to seek medical attention and to respond to the survey. Compared to older people, younger people are assumed to be more likely to have a moderate or severe reaction (henceforth referred to as severe reaction) [18, 20], but less likely to respond to a health survey [21, 22], and less likely to seek medical attention if they experience an AEFI. [23]. Table 1 defines variables in the simplified DAG, with parameters designed to generate data for each variable using Monte Carlo methods. Parameter values are chosen to describe a scenario for a low prevalence of severe reaction, low survey participation and a weak influence of severe reaction on both survey participation and MA. We subsequently used this as a reference scenario for PPA investigation. See the supplementary material for further details about how these parameters were used to generate the event probabilities in the data simulations, including the probability of a participant reporting MA.

To facilitate the investigation of how combinations of these factors influence signal detection, we simulated data for the reference scenario consisting of 50,000 hypothetical vaccine recipients representing accumulated safety data to date, and 12 investigation scenarios consisting of 4,000 hypothetical vaccine recipients representing a typical number of surveys issued to vaccine recipients in a two week investigation period.

In contrast with the reference scenario of a low probability for severe reaction (Low R), low survey participation (Low P), and a weak influence of severe reaction on survey participation and MA (Weak). We altered the value of four parameters outlined in Table 2 and generated 5,000 simulations for each investigation scenario. We varied the probability of severe reaction (*θ*) from low (Scenario 1-6) to high (Scenario 7-12) arbitrarily, in other words, under Scenarios 7-12 there’s a true change in the biological process compared with the reference scenario, i.e., a true increase in the risk of severe reaction as may plausibly occur due to a manufacturing issue with a particular vaccine batch. We varied the probability of survey participation (*η*) representing a change in the behaviour of vaccine recipients (Medium P for Scenario 2, 5, 8, 1 and High P for Scenario 3, 6, 9, 12). Finally, we varied the influence of severe reaction on both the probability of survey participation (*τ_sp_*) and the probability of MA (*τ_ma_*) (Strong Influence for Scenario 4-6, 10-12).

**Table 2:**
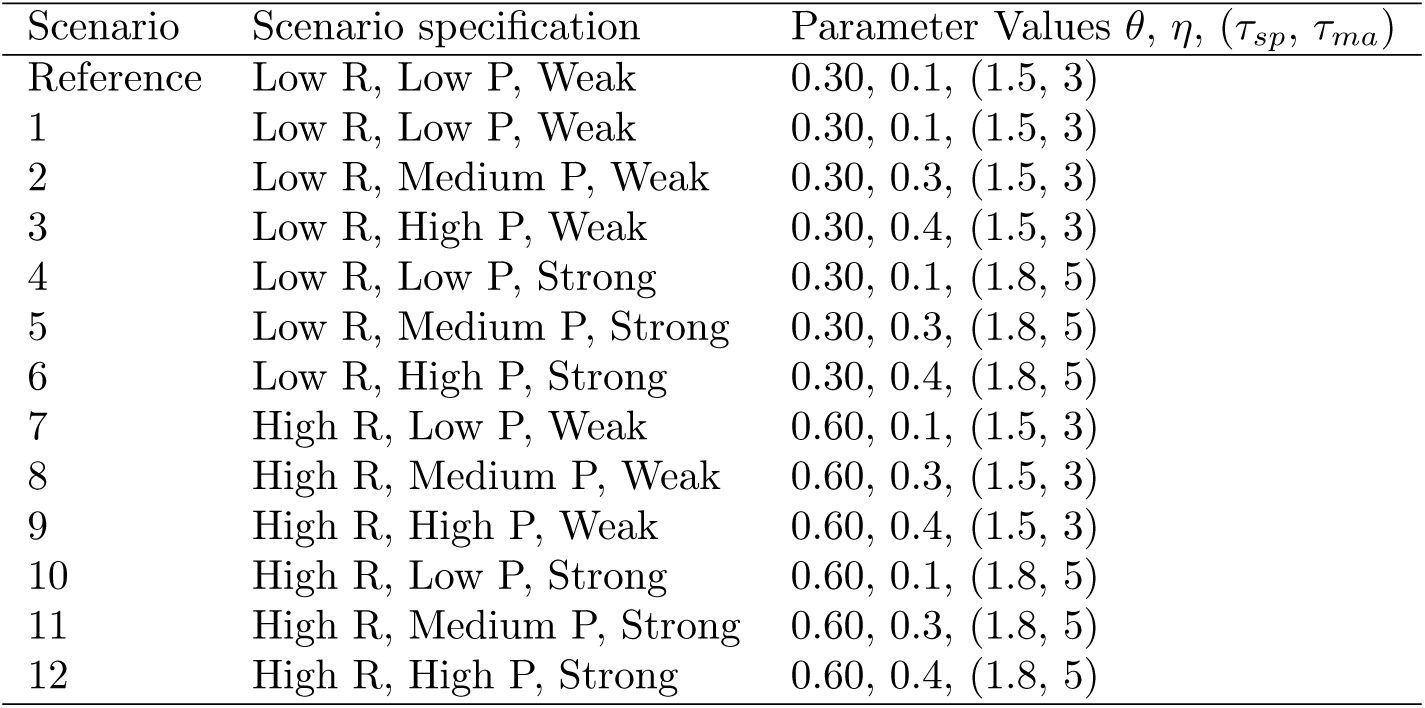
Definition of investigation scenarios. Scenarios vary with probability of severe reaction (Low or High R), probability of survey participation (Low, Medium or High P) and influence of severe reaction on the probability of survey participation and the probability of MA (Weak, Strong)

| Scenario | Scenario specification | Parameter Values $\theta, \eta, (\tau_{sp}, \tau_{ma})$ |
| --- | --- | --- |
| Reference | Low R, Low P, Weak | 0.30, 0.1, (1.5, 3) |
| 1 | Low R, Low P, Weak | 0.30, 0.1, (1.5, 3) |
| 2 | Low R, Medium P, Weak | 0.30, 0.3, (1.5, 3) |
| 3 | Low R, High P, Weak | 0.30, 0.4, (1.5, 3) |
| 4 | Low R, Low P, Strong | 0.30, 0.1, (1.8, 5) |
| 5 | Low R, Medium P, Strong | 0.30, 0.3, (1.8, 5) |
| 6 | Low R, High P, Strong | 0.30, 0.4, (1.8, 5) |
| 7 | High R, Low P, Weak | 0.60, 0.1, (1.5, 3) |
| 8 | High R, Medium P, Weak | 0.60, 0.3, (1.5, 3) |
| 9 | High R, High P, Weak | 0.60, 0.4, (1.5, 3) |
| 10 | High R, Low P, Strong | 0.60, 0.1, (1.8, 5) |
| 11 | High R, Medium P, Strong | 0.60, 0.3, (1.8, 5) |
| 12 | High R, High P, Strong | 0.60, 0.4, (1.8, 5) |

### PPA investigation

We paired the reference scenario with each investigation scenario, and assessed how likely the PPA method is likely to detect a signal under each investigation scenario, for which we present a histogram of where each simulation’s number of MAs under the investigation scenario sits as a percentile of the predicted probability distribution in Figure 4 and 5. We also presented here the percentage of simulations that generated a signal for each scenario by age group in green and red text. Figure 4 consists of the 6 investigation scenarios (1-6) where there is no change in the biological processes of interest compared with the reference scenario, and Figure 5 consists of those (Investigation Scenarios 7-12) where there is an increase compared with the Reference scenario (from Low R to High R).

**Figure 4:**
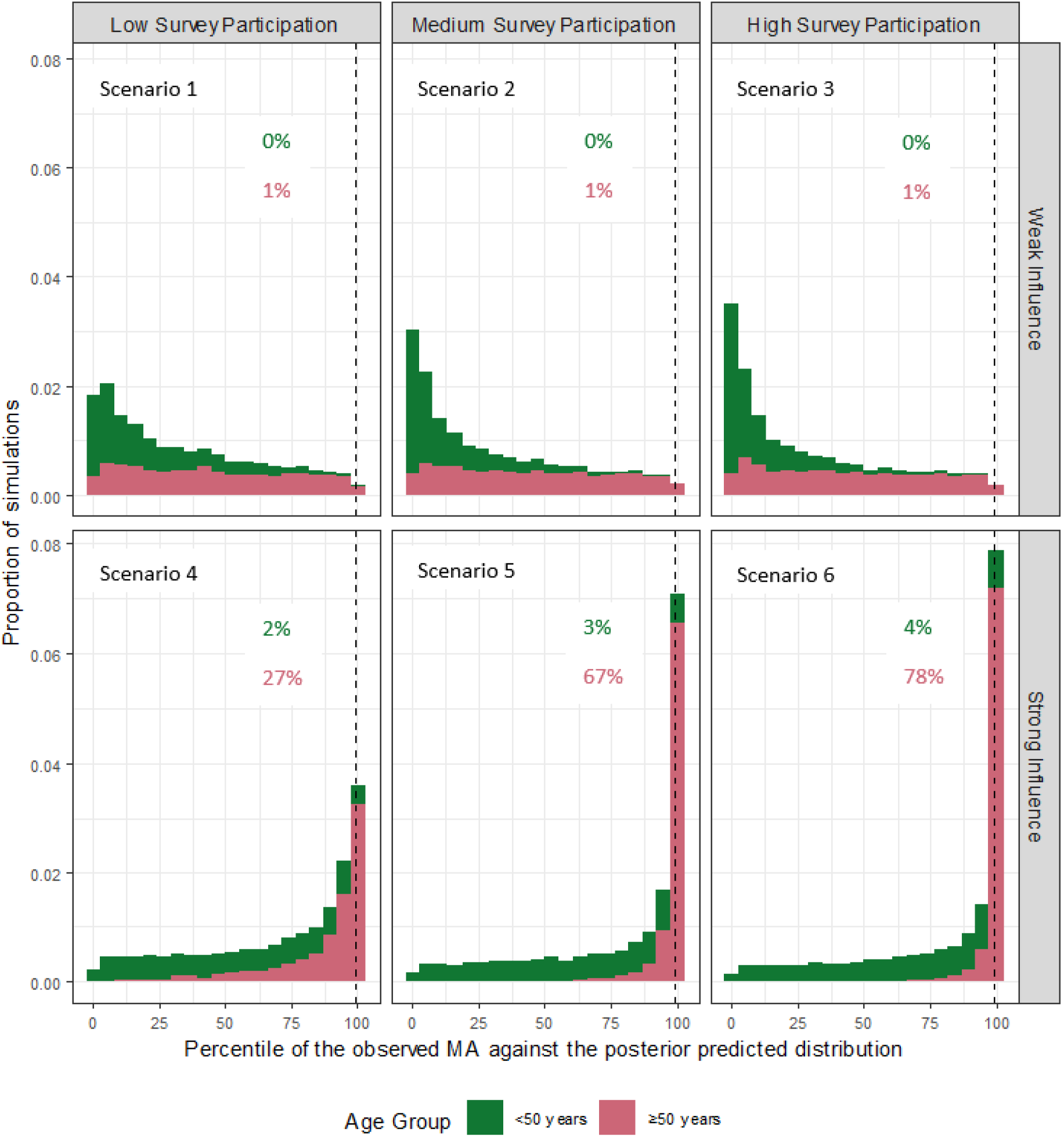
Low Probability of Severe Reaction in Investigation Scenarios, No change compared to the reference scenario. Signals are flagged when the number of MAs exceeds the 99th centile (dotted lines). The proportion of simulations which generated a signal within each scenario are represented as percentages within each panel in this figure. See Table 1 and 2 for simulation parameters used in each scenario.

**Figure 5:**
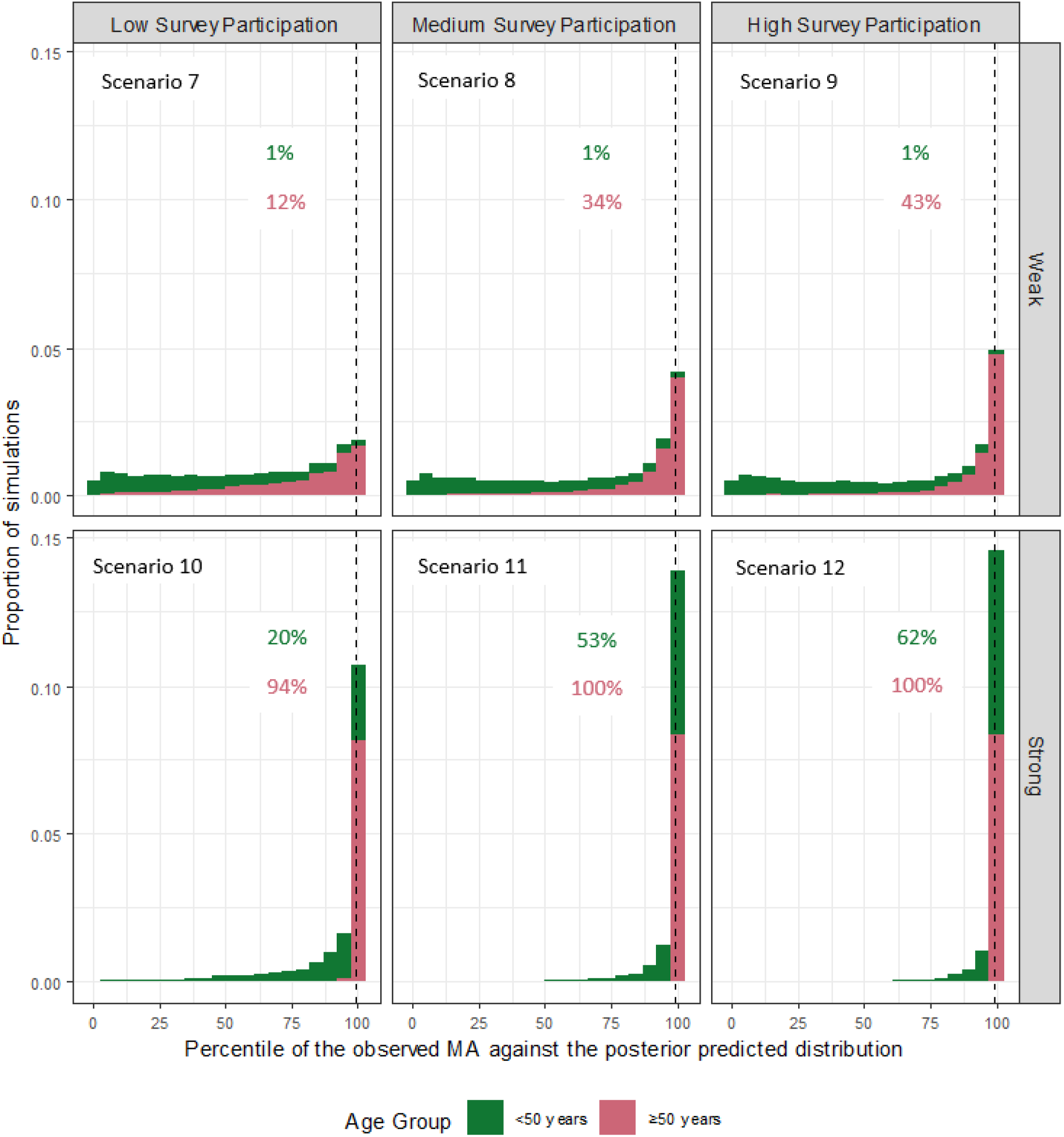
High Probability of Severe Reaction in Investigation Scenarios, Increase in biological processes compared to the reference scenario. Signals are flagged when the number of MAs exceeds the 99th centile (dotted lines). The proportion of simulations which generated a signal within each scenario are represented as percentages within each panel in this figure. See Table 1 and 2 for simulation parameters used in each scenario.

It is desirable for the PPA signal detection method to generate a safety signal in investigation scenarios that are set with a high prevalence (probability) of severe reaction (Figure 5) and not to generate a signal in scenarios set with a low prevalence of severe reaction (Figure 4). Any signals generated in simulations of high and low prevalence of reaction scenarios were therefore considered to be appropriate and inappropriate, respectively.

In low reaction prevalence (Low R) scenarios (Figure 4), signals were inappropriately generated in a moderately high proportion of scenarios involving the older age group when the influence of reactions on survey participation and MA was strong (27-78% of simulations), especially when survey participation was high. For the younger age group, only a low proportion of simulations resulted in an inappropriate signal generation (0-4% of simulations).

In high reaction prevalence (High R) scenarios (Figure 5), signals were appropriately generated in a high percentage of simulations of scenarios involving the older age group when the influence of reactions on survey participation and MA was strong (94-100% of simulations), regardless of the participation rate. The probability of appropriate signal generation was lower (1-62% of simulations) in the younger age group, and in the older age group when the influence of reactions on survey participation and MA was weak (12-43% of simulations).

## Discussion

Key challenges to vaccine safety monitoring are the low prevalence of AEFI, the relatively large number of vaccine recipients required to respond to safety surveys to ensure adequate sensitivity of the detection methods and the absence of information from vaccine recipients who do not participate in AEFI surveillance surveys. There is a need to balance sensitive signal detection methods that can detect true vaccine safety issues in a timely manner, while minimising the frequency of false detections. To understand this problem, we combined causal methods and statistical modelling, and used simulation to investigate how AEFI signal detection may be affected by biological and behavioural factors.

We used the novel PPA methodology based on a Bayesian logistic model of the probability of reporting medical attendance following an AEFI. Fundamentally, a PPA signal is generated by the AusVaxSafety monitoring system when the number of reported MAs for AEFI (for a specified vaccine) exceeds a threshold based on historically observed rates and the number of respondents in the current reporting period. There are two key reasons why a signal might occur. First, there might be a true increase in AEFI in the vaccinated population (i.e., the biological process of interest) which is reflected in increased reports of MA in the surveyed population. Second, in the absence of a true increase in AEFI, the survey respondents might be enriched with a subset of a population whose rate of reporting MA is higher; this could be due to changes in behaviour (such as media reports) and/or procedural processes (such as an age-related roll out of a vaccine) and independent of any change in the biological processes. This possibility adds to the practical challenge of detecting a true increase in AEFI and may trigger time-consuming case-series investigations by public health researchers.

A desirable safety signal detection system should be sensitive to changes in the true prevalence of severe vaccine reactions while being robust to variation in certain behavioural factors that might affect reporting, including the survey participation rate and the influence of reaction severity on the propensity to participate in the survey or to seek medical attention. However, we found evidence that our safety method could be sensitive to both behavioural factors. In high reaction prevalence investigation scenarios (Figure 5), signals failed to be generated in the younger age group if either the influence of reactions on survey participation and MA was weak or survey participation was low. In the older age group, signal generation was robust to low survey participation if the influence of reactions on survey participation and MA was strong. Conversely, in low reaction prevalence investigation scenarios (Figure 4), inappropriate signals can occur when there is a strong influence of reaction severity on survey participation and MA, especially when survey participation is high. An inflation of inappropriate or false signal detection may have a subsequent effect on investigative resources.

Inspection of the causal DAG (Figure 3), which underpinned the data generation for the simulation study, partly points to the explanation for the sensitivity of the signal detection method to changes in these behavioural factors. In essence, we wish to use data on reported MAs to make indirect inference about the (possibly changing) causal effect of vaccination on the prevalence of severe reactions. Severe reactions are only ascertained as reported medical attendances, which are conditionally dependent on both medical attendance occurring and survey participation. Independent of the influence of reaction severity, the probability of a MA is plausibly influenced by age and other factors. While the age of vaccine recipients can be measured and conditioned upon, these other factors are mostly unknown, unmeasured, and will therefore confound attempts to attribute any changes in MA to changes in the reactogenicity of the vaccine. Furthermore, changes in survey participation rates will also influence the rate of reported MA independent of any true increase in MA. If these changes are driven by factors other than changes in the age-distribution of vaccine recipients, there is a risk those factors will further confound the attribution of changes in reported MAs to changes in vaccine reactogenicity. Systematically monitoring survey response rates and survey participation behaviour might be used to determine the significance of an alerted signal and to improve the sensitivity of signal detection. When a signal has been alerted, it may be important to assess whether this is explainable by changes in survey participation behaviour, and this may inform the investigation and interpretation of the signal, *e.g.*, a signal alerted in the context of consistently high survey participation may have greater significance than a signal alerted in the context of varying survey participation behaviour.

Apart from the potential of changes in survey participation and propensity for MA to confound the attribution of changes in reported MAs to changes in vaccine reactogenicity, the PPA signal detection method is also sensitive to the quantity of data available at each analysis. Low rates of survey participation reduce the amount of data available to inform the parameters of the statistical model, and therefore reduce the certainty regarding the value of these parameters. As a result, even a moderately high frequency of reported MAs caused by a true increase in reactogenicity will be compatible with the statistical model (*i.e.*, the plausibly expected rate of reported MAs), so signals may not be generated even when the risk of severe reactions is high, thus it is desirable to increase survey participation by vaccine recipients. Promotion by immunisation providers at the time of vaccination may result in greater survey response rates.

The use of causal DAGs was useful for our study in several ways. The full DAG (Figure 2) depicts the problem domain of vaccine safety monitoring and thus facilitates communication among people from disparate disciplines, including medical experts, public health practitioners and statisticians. It provided a common starting point for simplification of the DAG and elicitation of the parameters and scenarios necessary for the data generation process. This is a necessary simplification of the real world but captures the important components of the complex target problem domain. The full DAG also serves as a knowledge base of the problem domain, which can support investigation of future research questions. Aided by the simplified DAG, we examined, by simulation, how changes in these non-biological factors might affect active surveillance data thus leading to a distorted interpretation of the biological process in the vaccinated population. Such simulations revealed how signal detection can be influenced by behavioural and procedural factors which affect survey participation and response but which cannot be gleaned from the survey data alone.

There are limitations to our simulation study. For simplicity, the only variable included in the PPA method was age, although other factors are known to drive biological, behavioural and procedural processes and thus affect how a detected signal should be interpreted. The AusVaxSafety PPA model is more complex and also accounts for gender, ethnicity, jurisdiction and co-morbidities, in addition to the age of the vaccine recipient. Also, we conducted our simulation study over two age groups when in reality, biological and behavioural factors may differ across age groups below the age of 50 years. Incorporating more realistic PPA models (with factors like gender and ethnicity, as well as greater granularity in the age groups) into a simulation study would be of interest in future analyses. Here we focused on the effect of differential survey responses among those who attend participating immunisation clinics. In addition, people attending participating immunisation clinics may be systematically different from those attending other immunisation services (*e.g.*, in remoteness). These differences may, in turn, affect survey response and healthcare seeking behaviours so reported AEFI rates may be over or underestimated. The current full DAG can be extended to depict this selection bias to reflect how the surveyed and responder population relates to the whole vaccinated population. While this may not impact the operating characteristics of our signal detection, future work will attempt to account for this selection bias in AEFI rate estimation by accounting for the type of clinic in the statistical model. Also, we did not differentiate between seeking phone advice for AEFI versus GP or hospital attendance, nor did we consider detection via passive (spontaneously volunteered) rather than active (solicited) AEFI reports. Greater granularity for these factors may be incorporated into future models.

## Author contributions

YW, TS and JM conceptualised this study. Causal modelling, simulation, and statistical analysis were devised and conducted by ET, YW, MD and SB. YH, CG, LL and TS provided domain expertise. All authors contributed to the interpretation of results and the writing of the manuscript. All authors approve of distribution of this manuscript.

## Supporting information

Supplementary Dictionary

## Data Availability

Data simulation code is available on Github. See link below and also within the manuscript

https://www.github.com/ECSTay/AVSCausalModel

## Acknowledgement

This project is supported by the Australian Government Department of Health and Aged Care. Dr Lucy Deng contributed her clinical expertise in constructing the full causal model for this study.

