## Supplementary Material for "Applying causal inference and Bayesian statistics to understanding vaccine safety signals — a simulation study"

### Supplementary Material - Data Simulation Parameters

The following table describes the parameters used to generate the baseline scenario for a low prevalence of severe reaction, low survey participation and small influence of severe reaction on both survey participation and seeking medical attention (MA).

Table 1: Data simulation parameters

| Parameters | Definition | Values |
|---|---|---|
| $\theta$ | prevalence of severe reaction in <50y age group | 0.3 |
| $\epsilon$ | relative reduction of prevalence of severe reaction in ≥50y age group | 0.3333 |
| $\eta$ | probability of survey participation in the <50y age group with mild reactions | 0.1 |
| $\tau_{sp}$ | relative increase of survey participation due to severe reactions | 0.5 |
| $\mu_{sp}$ | relative increase of survey participation due to older age | 0.35 |
| $\phi$ | MA in the <50y age group with mild reactions | 0.01 |
| $\tau_{ma}$ | relative increase of MA due to severe reactions | 2 |
| $\mu_{ma}$ | relative increase of MA due to older age | 4 |

Using these parameters, we obtained event probabilities in each subgroup. The open source code used to generate the simulated data for each scenario is available: www.github.com/ECSTay/AVSCausalModel

Table 2: Generation of event probabilities within the scenarios

| | Prevalence of Moderate - Severe Reaction |
|---|---|
| <50y | $\theta$ |
| $\geq 50y$ | $\theta$ * (1 - $\epsilon$) |
| | **Probability of Survey Participation** |
| <50y, mild reaction | $\eta$ |
| <50y, severe reaction | $\eta$ * ( 1 + $\tau_{sp}$) |
| ≥50y, mild reaction | $\eta$ * (1 + $\mu_{sp}$) |
| ≥50y, severe reaction | $\eta$ * ( 1 + $\mu_{sp}$) $*$ (1+$\tau_{sp}$) |
| | **Probability of Seeking Medical Attention** |
| <50y, mild reaction | $\phi$ |
| <50y, severe reaction | $\phi$ * ( 1 + $\tau_{ma}$) |
| ≥50y, mild reaction | $\phi$ * (1 + $\mu_{ma}$) |
| ≥50y, severe reaction | $\phi$ * ( 1 + $\mu_{ma}$) $*$ (1+$\tau_{ma}$) |
